# Comparative Inference of Gene-Specific and Global Gene Regulatory Network Methodologies for Biomarker Assessment in Lung Cancer

**DOI:** 10.1101/2024.10.23.24315971

**Authors:** Supriya Mandal, Subhrapratim Nath, Surama Biswas

## Abstract

Understanding biomarker transitions from normal physiological states to malignancy requires moving beyond differential expression toward regulatory network level characterization. This study aims to advance gene regulatory network (GRN) based biomarker transition methodologies by capturing condition specific regulatory rewiring in lung cancer (LC). Two independent LC gene expression datasets were harmonized, integrated, and normalized for unified network inference. A robust gene set was obtained through the intersection of simulated annealing based selection, variance based screening, and volcano plot analysis. Regulatory interactions were inferred separately for normal and disease states using coexpression networks, along with our proposed Focused Random Forest (FRF) based GRNs, and Global Dual Random Forest (GDRF) framework. A comparative analysis revealed widespread disruption of coordinated gene expression and substantial regulatory remodeling during disease progression. Centrality measures identified CLDN18, CPB2, GJB2, MT1M, MMP12, WIF1, and GREM1 as key biomarker drivers. Directed GRNs demonstrated gene specific phase transitions through shifts in regulator target relationships. GDRF analysis revealed a transition in regulatory dominance from GJB2 and GREM1 in the normal state to CLDN18 and CPB2 in LC. The novelty of this work lies in the unified integration of local and global Random Forest based GRN inference to quantify biomarker phase shifts through directed regulatory remodeling.

## 1. Introduction

Analytical frameworks that move beyond static expression-level indicators are required to elucidate the transition of molecular systems from normal physiological states to cancer. The regulatory circuitry that controls cellular state transitions is not well understood by differential gene expression analysis, despite its widespread use in identifying genes linked to disease [1]. Biomarkers can be viewed in terms of their functional roles within dynamic networks rather than isolated expression variations thanks to gene regulatory networks (GRNs), which provide a potent systems-level paradigm for modeling gene–gene relationships and regulatory effects [2]. GRN-based biomarker transition study specifically examines how the regulatory significance, connectivity, and interaction landscapes of genes change between healthy and diseased conditions [3]. By capturing coordinated gene interactions rather than isolated expression changes, GRNs facilitate the identification of disease biomarkers. Collective regulatory disturbances drive the genesis and progression of complex illnesses like hepatocellular carcinoma. Early detection of crucial transitions is made possible by the observation of dynamic network rewiring across illness phases, made possible by temporal GRNs. In contrast to traditional gene-level techniques, network-based research thus identifies dynamic network biomarkers (DNBs) with enhanced resilience and biological interpretability. [4].

LC is one of the leading and widespread forms of cancer globally. It accounts for 12.3% of all cancer diagnoses. Between 2000 and 2022, the number of new LC cases increased from 1.2 million to more than 2.4 million. LC is, by historical data, the most frequent cancer in males, with a high morbidity and mortality in both genders. Demographic and geographic differences significantly impact its occurrence. LC is highly preventable by reducing smoking habits and taking sustainable measures to improve air quality. Though many preventative measures have already been implemented, the risk never goes back to normal, and fatal conditions persist with a 90% patient death rate [5]. After the COVID-19 pandemic, patients with LC should have poorer prognoses, as they have a higher risk of death from the virus and may experience severe consequences through contamination. Accordingly, a few recent studies claimed that individuals with COVID-19 who developed LC had far worse death rates than the COVID-19 community as a whole [6 - 7]. Current research focus on LC is to study the pathology, early detection, and prognosis mechanisms, and understand how different environmental and genetic variables impact the disease [8].

Recent advancements in LC research have significantly improved the understanding of the disease through the application of genomic and proteomic technologies. These technologies offer comprehensive insights beyond single-gene studies, enhance our knowledge of lung carcinogenesis, and have the potential to improve diagnosis, prognosis, and patient management [9]. Genomic research has focused on understanding the histologic differentiation and progression of LC, revealing critical genetic alterations that drive the disease [10]. The emergence of individualized diagnostics and treatments has revolutionized LC care with specific therapies targeting genetic alterations. Such targeted alterations are found in Estimated Glomerular Filtration Rate (EGFR)-mutant adenocarcinoma that leads to more effective treatments compared to traditional chemotherapy [11]. Genomic testing has become integral in identifying mutations and resistance mechanisms in LC, aiding in the development of precision medicine approaches that deliver treatments to individual genetic profiles. This testing is now a part of national guidelines due to its demonstrated benefits in improving patient outcomes [12]. Additionally, research on intra-tumor heterogeneity (ITH) has demonstrated that transcriptome variability inside tumors plays a role in therapeutic resistance and immune evasion. Researchers have connected gene expression patterns to tumor expansion and metastasis by examining coupled whole-exome and RNA sequencing data, underscoring the intricate interaction between the transcriptome and genome in the development of LC [13].

GRN reconstruction is pivotal in systems biology, providing insights into gene interactions and regulatory mechanisms within cells. Utilizing correlation and regression analyses of multifactorial perturbation data, a method decomposes the GRN inference problem into different regression tasks, predicting target gene expression levels from potential regulator genes. This approach, integrating correlation coefficients and sum of squared residuals, has shown promising results in accurately estimating gene regulations [14]. GRNs serve as comprehensive maps reflecting genetic and epigenetic influences, aiding in understanding the functions of individual molecules and overall cellular organization [15]. Differential GRNs capture variable gene expression under different conditions, crucial for studying development, organ formation, and disease progression through high-throughput sequencing data and gene-gene expression correlation analysis [16].

Random Forest is an ensemble learning technique that creates several decision trees on randomized data subsets and aggregates their outputs to produce precise and reliable predictions, even in high-dimensional and noisy environments [17]. It is a dependable option for GRN inference from gene expression data because of its capacity to manage missing values, represent nonlinear interactions, and minimize overfitting. Random Forest’s strong prediction accuracy in categorizing network-based gene characteristics highlights its efficacy in finding important regulatory genes and validates its dependability for biomarker discovery and GRN inference [18]. A network-guided Random Forest method that integrates gene network data into feature sampling for gene expression analysis is assessed in a study. As shown in Cancer Genome Atlas (TCGA) breast cancer datasets, this approach improves disease gene discovery when genes form coherent network modules, even though it does not increase disease prediction accuracy over standard Random Forest. However, when network assumptions are broken, it may introduce spurious selections [19]. Decision tree-based models, which are the ancestors of Random Forest, offer an interpretable basis for GRN inference. In order to demonstrate improved accuracy and extensibility in GRN reconstruction, a recent research work proposes an intriguing dual-decision tree framework (BFMDDT) that integrates multi-type gene expression datasets by independently modeling regulator-to-target and target-to-regulator relationships and combining their feature importance rankings [20].

GRN analysis is one of the most important means of identifying biomarkers for various diseases. The spectrum includes sepsis and cancer. In order to improve the sensitivity and specificity of sepsis diagnostics, researchers have discovered new miRNA biomarkers linked to the early diagnosis of sepsis by concentrating on microRNA (miRNA) regulatory networks [21]. Comparably, GRN-based methods have shown important roles in LC and ovarian cancer biomarker detection. In a pathway linked to these tumors, for instance, the overexpression of the gene RAD51AP1 is actually associated with a lower overall survival rate. Gene suppression research supports this conclusion, which emphasizes the function of RAD51AP1 in the growth of cancer and its potential as a predictive biomarker [22]. Certain gene signatures that can predict patient prognosis have been found in bladder cancer through the development of a competitive endogenous RNA (ceRNA) regulatory network, including lncRNA-miRNA-mRNA interactions. This ceRNA network, developed using data from The TCGA, has provided insights into the molecular mechanisms underlying bladder cancer and has established a model for prognostic prediction [23].

Network-based approaches have significantly advanced the identification and understanding of biomarkers for cancer, which enables researchers to explore gene interactions and regulatory mechanisms of the disease. These methodologies integrate human transcriptome and interactome data and prioritize genes in cancer-specific networks that lead to the identification of cancer genes crucial for tumorigenesis and progression. These genes, showing extensive perturbations, contribute to accurate tumor diagnosis and patient survival prediction [24]. Utilizing microarray gene expression profiles and protein-protein interaction information, network-based biomarkers have been developed to explore lung carcinogenesis mechanisms, highlighting pathways and potential therapeutic targets [25]. The Gene Sub-Network-based Feature Selection (GSNFS) approach, incorporating novel searching and scoring algorithms which has further refined subnetwork identification, improved classification performance and outperforms across datasets as compared to traditional greedy algorithms [26]. In lung adenocarcinoma (LUAD), Weighted Gene Co-expression Network Analysis (WGCNA) has identified key gene modules and hub genes related to cancer stem cell features, with prognostic signatures constructed from these hub genes demonstrating high predictive value [27]. Moreover, transcriptional regulatory networks have been constructed to identify dysregulated transcription factors in LC, revealing co-expressed and co-regulated patterns linked to critical biological processes and cancer hallmarks. These dysregulated transcription factors, validated through enrichment analysis and survival correlation studies, offer potential for early diagnosis and personalized therapies [28].

The goal of this paper is to develop GRN-based biomarker transition approaches for a systems-level understanding of disease development. Characterizing the shift from normal physiological states to malignancy, especially in complex disorders like LC, requires capturing regulatory rewiring rather than isolated expression alterations. In order to facilitate collaborative and consistent network inference, two separate seperate LC gene expression datasets were combined via strict gene-level harmonization. This integration makes it easier to compare regulation modelling across physiological states and prevents fragmented findings from dataset-specific analysis. Three complementary gene selection techniques—simulated annealing-based approach [29], a variance-based screening, and volcano plot analysis—were used following normalization and data modification. A strong group of 11 important genes, including CLDN18, CPB2, GJB2, MMP12, WIF1, AGER, and GREM1, was produced by the intersection of these methods and was used as potential biomarkers for further investigation. Multi-scale regulatory dynamics were captured using both associative and causal network modelling frameworks. In order to discover global rewiring tendencies and create baseline association patterns, co-expression networks were initially built for both healthy and pathological states. A targeted modelling approach was then utilized to infer Random Forest-based GRNs to identify directed regulatory connections among the discovered genes. Quantitative evaluation of regulatory phase shifts between normal and LC states was made possible by a comparative examination of network topology utilizing several centrality measures. Our proposed approach, titled Global Dual Random Forest (GDRF) to infer GRN, in which each identified gene through the gene selection step is alternately treated as a target and as a regulator, extracting up to three genome-wide regulators and targets, respectively, and integrating both views into a unified adjacency matrix. This extends prior decision-tree–based dual inference [20] to a more stable and scalable Random Forest paradigm. Centrality analysis of GDRF–derived GRNs identified CLDN18, CPB2, GJB2, and GREM1 as key regulatory drivers underlying biomarker transitions. Notably, GDRF centrality profiles revealed a transition in regulatory dominance from GJB2 and GREM1 in the normal state to CLDN18 and CPB2 in lung cancer, indicating a reorganization of control within the regulatory landscape. The novelty of the proposed approach lies in the unified integration of local and global Random Forest–based GRN inference, enabling centrality-guided quantification of biomarker phase shifts through directed regulatory remodeling. The methodical assessment of how important biomarkers both impact and are impacted by the larger regulatory environment was made possible. The highlights and key contributions are as listed below:

- Integrated co-expression and Random Forest–based causal GRNs to characterize biomarker transitions, using centrality for quantitative assessment and regulator–target rewiring for qualitative analysis of disease progression.
- Novel dual Random Forest GRN inference treating each biomarker gene as both target and regulator to identify top genome-wide regulators and targets, integrated into a unified GDRF network.
- Biomarker transitions in LC are demonstrated at the gene-specific and genome-wide levels. GDRF centrality profiles revealed a transition in regulatory dominance from GJB2 and GREM1 in the normal state to CLDN18 and CPB2 in LC, indicating a reorganization of control within the regulatory landscape.

## 2. Methodologies

This section presents a complete study of LC genomics. It integrates two gene expression datasets, perform preprocessing steps and transposition, next applies three statistical and/or AI-ML methods for gene selection, identifies overlaps among the selected genes, infers GRNs among the genes associated with LC in healthy and diseased states, obtains centrality measures of the two GRNs, and presents a comparative study of network biomarker transitions from healthy to diseased states.

### A. Data Collection and Pre-Processing Procedures

Gene expression datasets reflecting differential activity between healthy and diseased lung tissues were obtained from publicly available repositories. Two independent LC gene expression datasets were selected to enable integrative analysis.

In the original datasets, probe identifiers were provided as row headers and patient samples as columns. Probe IDs were mapped to official gene symbols using platform-specific family description files. Probes without valid gene symbol annotations were removed. When multiple probes mapped to the same gene symbol, their expression values were averaged to generate a single representative expression profile per gene.

Each dataset was independently cleaned and sorted based on gene symbols to ensure consistent ordering. Only the intersecting set of genes common to both datasets was retained. The resulting expression matrices were then horizontally concatenated across samples to construct an integrated gene expression matrix. This strategy preserves shared biological signals while minimizing dataset-specific inconsistencies. The integrated dataset was subsequently normalized and transposed to obtain a samples × genes matrix suitable for downstream statistical analysis and GRN inference.

### B. Gene selection procedures

Three separate approaches have been used in parallel to identify a small number of significant genes from the massive dataset: 1) a recent optimization method of gene selection known as Differential Gene Expression based Simulated Annealing (DGESA) [29], 2) screening of genes based on variance and 3) a Volcano plot method that applied statistical basis of significant gene identification.

#### 1) Selection using a Simulated Annealing-based Algorithm

A recent algorithm, called DGESA [29], has been implemented here for significant gene selection of LC. DGESA is based on the famous meta-heuristics algorithm, called Simulated Annealing (SA) [30], where a concept of metallurgy referred to as annealing is simulated to provide heuristic information traverse and assess the points in a search space of an optimization process against a properly designed objective function. In DGESA, this objective function is formulated in an efficient way (see Eq. 1) to maximize the cumulative mean difference of differential expression profiles (in healthy vs. LC samples) across all genes in a set (i.e. candidate solution comprising a set of gene indices selected from the dataset). DGESA starts with setting a few algorithmic parameters like temperature (T) set to a very high value, a cooling rate (C), and a randomly selected set of gene index into the initial candidate solution (s) of a fixed length. The solution s is evaluated by Eq. 1 and undergone few iterations. In each iteration, few neighborhood search steps are followed by a temperature reduction step using Eq. 2. In a neighborhood search step, a new solution *s*′ is generated from s using perturbation. If the objective function value of *s*′ is better than that of s, *s*′ replaces s, called a movement. Otherwise, *s*′ has a probability (p) dependent scope to replace s. DGESA terminates if the maximum of iterations is exceeded or if the objective function value of successive candidate solutions achieves convergence.

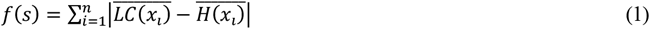

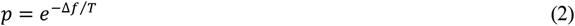

Here, *f*(*s*) is the objective function value calculated upon 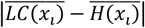 is the absolute value of the quantity, 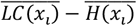 where 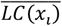 is the mean of all LC expressions of the current gene in *s* and 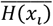 is the mean of all healthy expression values of the same gene.

#### 2) Selection by variance

Variance-based gene identification is a crucial step in understanding the differential expression of genes across samples. The variance of each gene’s expression levels is calculated to measure how much these levels deviate from their mean value. This involves computing the mean expression level for each gene (see Eq. 3) and then determining the variance using Eq. 4, which quantifies the spread of expression values around this mean. Genes are then sorted in descending order based on their variance. Those with the highest variance are selected for further analysis, as they are likely to exhibit significant differences in expression between healthy and LC tissues. The genes with the most variance, which may be the most biologically significant for differentiating between the various tissue types, are kept front and center by this approach.

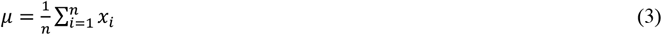

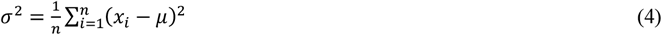

Here, *μ* is the mean expression value, *n* is the total number of healthy and diseased samples, respectively. *x*_*i*_ is the expression value of the i-th sample of the current gene and *σ*^2^ is the computed variance of the current gene across all healthy and diseased samples.

#### 3) Selection through Volcano Plot

For the identification of LC genes, a volcano plot has been generated here. In genetics and genomics, a volcano plot is a kind of scatter plot that is used to show the findings of a differential expression analysis. With a large number of genes or transcripts at the base and a smaller number of highly differentially expressed genes at the top, the plot is shaped like a volcano, hence the name “volcano plot.” The y-axis shows the statistical significance of the difference, which is shown as a −log10 (p-value), whereas the x-axis usually shows the log2 fold change of gene expression between two conditions (e.g., LC and healthy states). It is simple to determine which genes are most differentially expressed, as those with substantial fold changes and strong statistical significance are located in the upper left and right corners of the plot, above the predefined thresholds. Genes that are strongly up- or down-regulated under various situations can be found using volcano plots.

### C. GRN Inference

Three complementary GRN inference approaches were used to describe regulatory interactions among the discovered genes linked to LC. These methods record gene-gene connections at several levels, from directed, causal regulatory dependencies at both gene-specific and genome-wide scales to associative co-expression patterns. When combined, they allow for a thorough evaluation of network rewiring between healthy and pathological states as well as regulatory organization.

#### 1) Co-expression Network Construction

Using the selected genes, GRNs have been constructed for both healthy and LC tissue samples through a co-expression model. This process begins with a correlation analysis, where Pearson correlation coefficients are calculated (see Eq. 5) for each pair of genes to assess the strength and direction of their linear relationships. To define edges in the networks and suggest possible regulatory relationships between the genes, significant correlations are found. Network analysis methods are then used to show these GRNs, revealing the intricate network of interactions and regulatory processes. Differences in gene connections and regulatory mechanisms between healthy and LC tissues can be found by comparing their GRNs. This information can be used to understand how gene regulation changes in disease states.

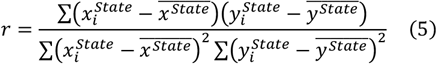

Here, 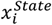 or 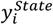 is an element in gene expression profile *x*^*State*^ or *y*^*State*^ in a certain *State*, whereas 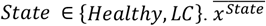 and 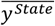 are the means of *x*^*State*^ or *y*^*State*^ respectively.

#### 2) Focused Random Forest (FRF) Model

A supervised, tree-based approach for deriving directed GRNs from gene expression data is called Focused Random Forest (FRF). By limiting network inference to a predetermined collection of biologically important genes, FRF enables targeted modelling of regulatory connections between potential biomarkers as opposed to genome-wide variables. In high-dimensional and noisy expression contexts, this architecture increases robustness, decreases computational complexity, and improves interpretability. Every gene in the chosen list is repeatedly treated as a target gene for a particular physiological state (healthy or ill), with the remaining genes acting as potential regulators. The candidate regulators are used as input features to train a Random Forest regression model to predict the target gene’s expression. Each regulator’s impact on the target gene is shown by the feature importance scores derived from the training model, which are interpreted as regulatory strengths. From each regulator to the goal, this procedure produces a directed, weighted edge. The distribution of significance scores in both healthy and diseased samples is used to calculate a data-driven threshold that separates strong regulatory interactions from weak or bogus ones. To create a filtered GRN, regulatory edges with significance values higher than this cut-off are kept. To quantify shifts in regulatory influence and information flow between physiological states, the resulting networks are examined using a variety of centrality metrics. FRF enables the systematic identification of regulatory rewiring and biomarker transitions associated with disease development by inferring GRNs independently for both healthy and diseased situations.

##### ALGORITHM 1

**FRF()**

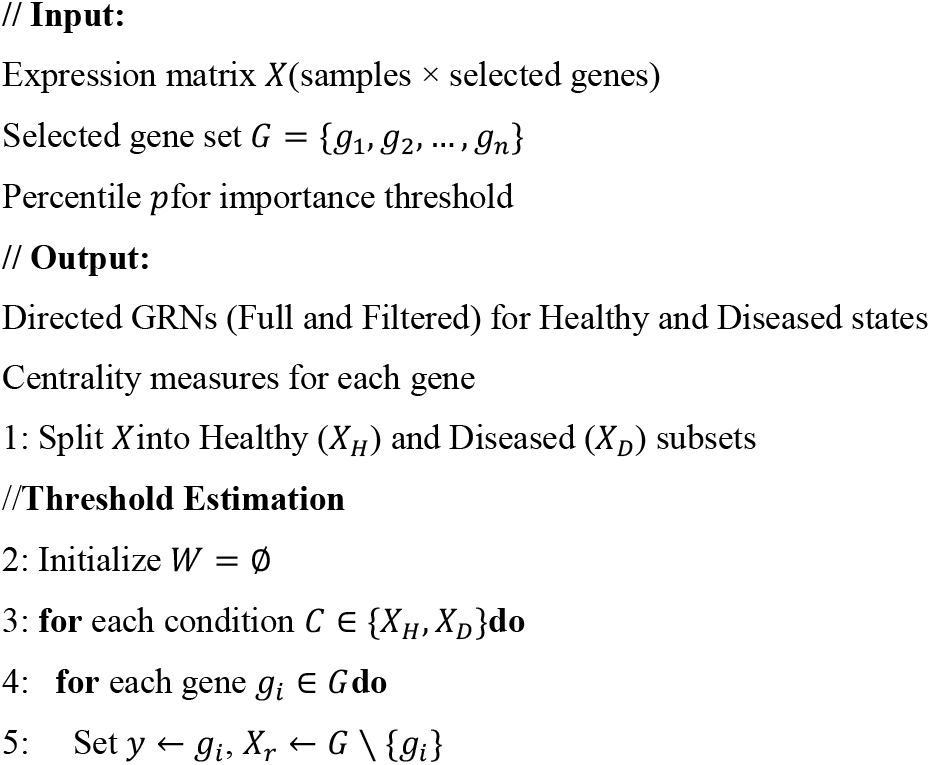
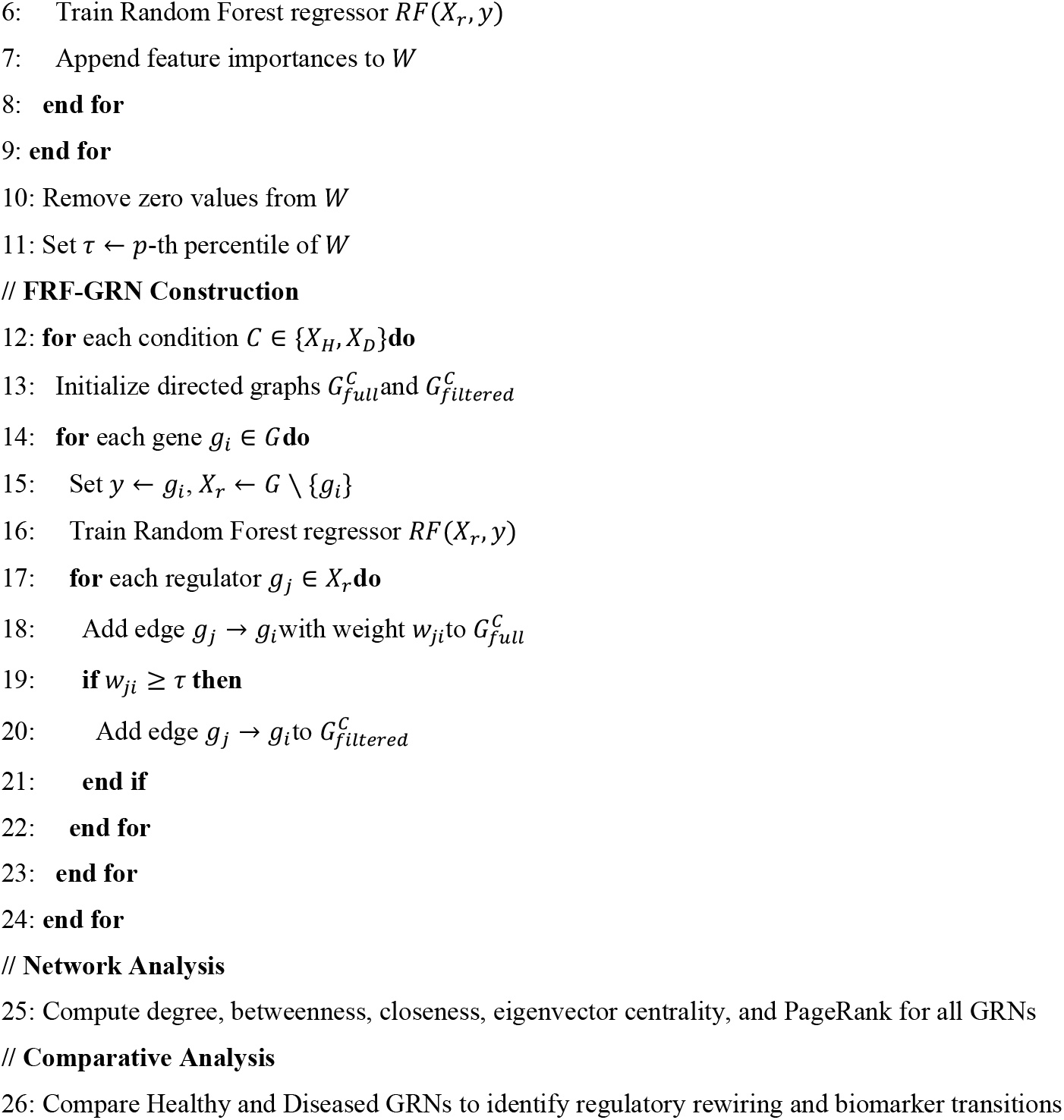

#### 3) GDRF Model

A GDRF-based GRN inference approach is used to identify genome-wide regulatory impacts linked to the selected biomarkers. To infer directed regulatory interactions from both regulator-to-target and target-to-regulator viewpoints, two complementary Random Forest regression models are built inside this framework. Based on Random Forest feature relevance scores, two weighted edge lists were independently constructed for each physiological state, one indicating potential regulators for each target gene and the other identifying potential targets regulated by each gene. To guarantee consistency among models, regulator and target gene symbols were standardized before integration, and significance values were cleaned and normalized. In order to reduce directional bias and increase robustness, the outputs of both models were then combined by creating a weighted adjacency matrix that incorporates regulatory evidence from both directions. The inferred regulatory strength from a regulator gene to a target gene is represented by each matrix entry. A directed GRN, in which nodes represent genes and weighted edges indicate inferred regulatory relationships, was then built using the resulting adjacency matrix. The found biomarkers may be examined within their larger genome-wide regulatory context by our dual modeling and integration approach, which also makes it possible to systematically characterize global regulatory rewiring.

##### ALGORITHM 2

**GDRF()**

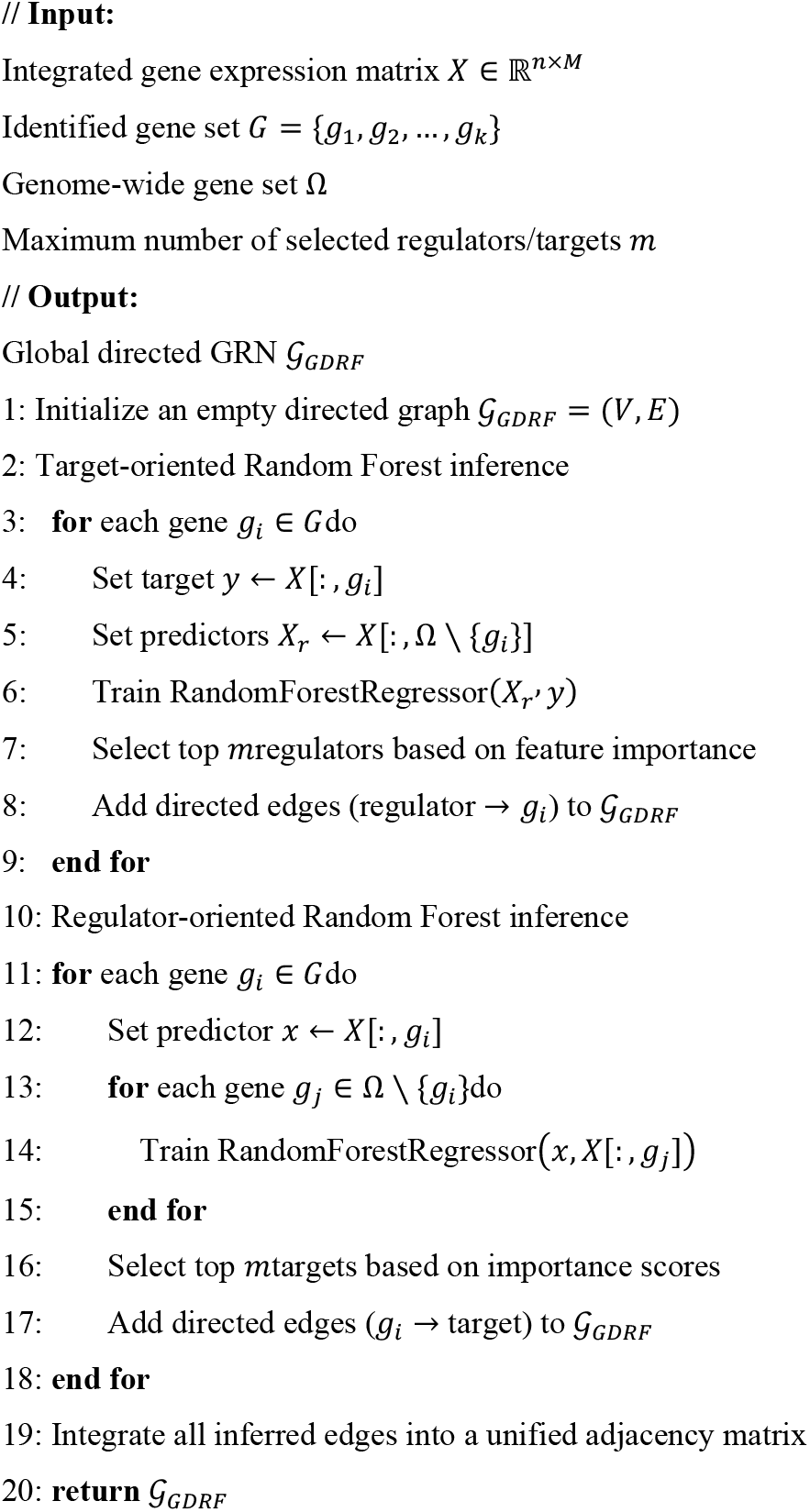

### D. Centrality Analysis

Centrality measures are metrics used to identify the most important nodes within a network. Here, it is performed on the differential GRNs obtained from the FRF model to determine each gene’s regulatory importance. Centrality measures that are considered here include:

1. **Degree Centrality**: This determines each gene’s immediate influence within the network by counting the number of direct connections it has.
2. **Closeness Centrality**: This measures a gene’s overall reach by calculating the reciprocal of the average shortest path length from a gene to every other gene. It evaluates how rapidly a gene can affect the entire network.
3. **Betweenness Centrality**: This highlights a gene’s function in promoting contacts by calculating the frequency with which it functions as a bridge within the network based on the number of shortest pathways between gene pairs that go through it.

Key regulatory genes with notable influence, reach, and bridging roles in the GRNs of healthy and sick tissues are found by computing these centrality metrics, which help identify prospective treatment targets and biomarkers.

### E Comparative Analysis between Healthy and Diseased Network Biomarkers

To find notable variations in gene regulation, the comparative analysis compares the centrality measures of genes in healthy and diseased GRNs. Key regulatory genes are identified in this method, particularly those whose centrality in the diseased network is high and whose centrality in the healthy network has changed noticeably. These genes are emphasized as possible important disease regulators. The structure and connectivity patterns of the GRNs are also investigated by comparing the overall network architecture. By exposing changes in network structure and interactions, this aids in the understanding of how LC affects gene regulation. Important network biomarkers can be found using this comparative study, which may reveal targets for therapeutic intervention as well as insights into the underlying mechanisms of LC. All steps of the methodology are comprehensively summarized in Fig. 1.

**Fig. 1.**
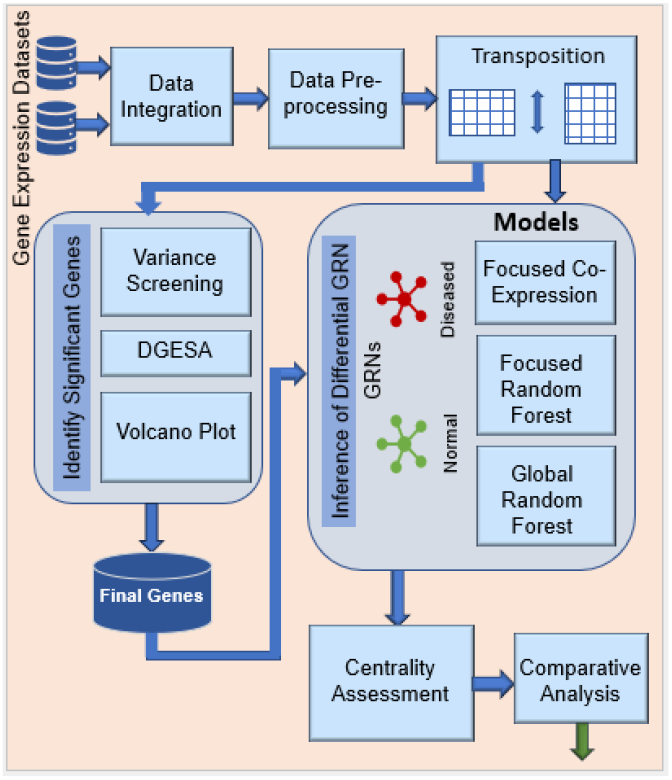
Overview of the proposed GRN-based biomarker transition framework: The workflow integrates gene expression datasets, identifies key genes, and infers co-expression, focused, and global Random Forest–based GRNs to analyze regulatory rewiring between healthy and diseased states.

## 3. Results

The results of integrated data analysis, gene selection, and GRN inference applied to LC are shown in the Results section. In order to clarify biomarker transitions, it highlights the identified important genes and methodically contrasts network topologies and regulatory changes between healthy and sick states.

### A Data Acquisition and Computational Resources

From the public database of National Center for Biotechnology Information (NCBI), Gene Expression Omnibus (GEO) [31-32], two separate gene expression datasets for non-small cell LC (NSCLC), named GSE18842 [33] and GSE19804 [34], were collected. In order to facilitate collaborative regulatory analysis, the datasets were integrated into a single unified matrix: first, platform annotation files were used for probe-to-gene symbol mapping. The rows with invalid/null gene symbols were excluded. To create a single gene-level profile for several probes representing the same genes, average expression levels were calculated. Only the genes that are common to both datasets were kept after each dataset was sorted by gene symbols. After that, horizontal concatenation was used to combine the aligned datasets across samples (see Supplements Table 1 for distribution of the number of genes and samples in different stages). To guarantee uniformity among samples, the integrated dataset was applied preprocessing like normalization between [0 - 1] and transposition. Robust gene selection, comparative network modeling, and GRN-based biomarker transition analysis within a common regulatory context were made possible by this unified preprocessing approach. Standard scientific libraries and Python 3.12.2 were used for all computational analyses (see Supplements Table 2 for details).

**Table I.** Common Genes Identified by Three Gene Selection Methods in the Integrated Dataset.

|  |  |  |
| --- | --- | --- |
| MMP12 [35] | GJB2 [39] | MMP1 [43] |
| WIF1 [36] | CPB2 [40] | GKN2 [44] |
| AGER [37] | TMEM100 [41] | CLDN18 [45] |
| GREM1 [38] | MT1M [42] |  |

### B. Critical Genes Underlying LC Pathogenesis

To identify robust genes associated with LC, three complementary gene selection strategies were applied to the integrated gene expression dataset: a simulated annealing–based optimization approach (DGESA) [29], variance-based screening, and volcano plot–based differential expression analysis. These methods collectively capture optimization-driven discriminability, global expression variability, and statistical significance. To ensure a fair and meaningful comparison across methods, threshold parameters for the variance-based and volcano plot–based approaches were carefully tuned to satisfy two objectives: (i) to obtain gene sets comparable in size to the SA-derived subset, and (ii) to retain a non-zero yet limited overlap among the methods. Using a variance threshold of 0.04, the variance-based approach yielded 43 highly variable genes, emphasizing genes with substantial expression fluctuations across samples, although this method does not explicitly account for class-specific separation. The overall outcomes of the three gene selection strategies are illustrated in Fig. 2 (Panels A–C).

**Fig. 2.**
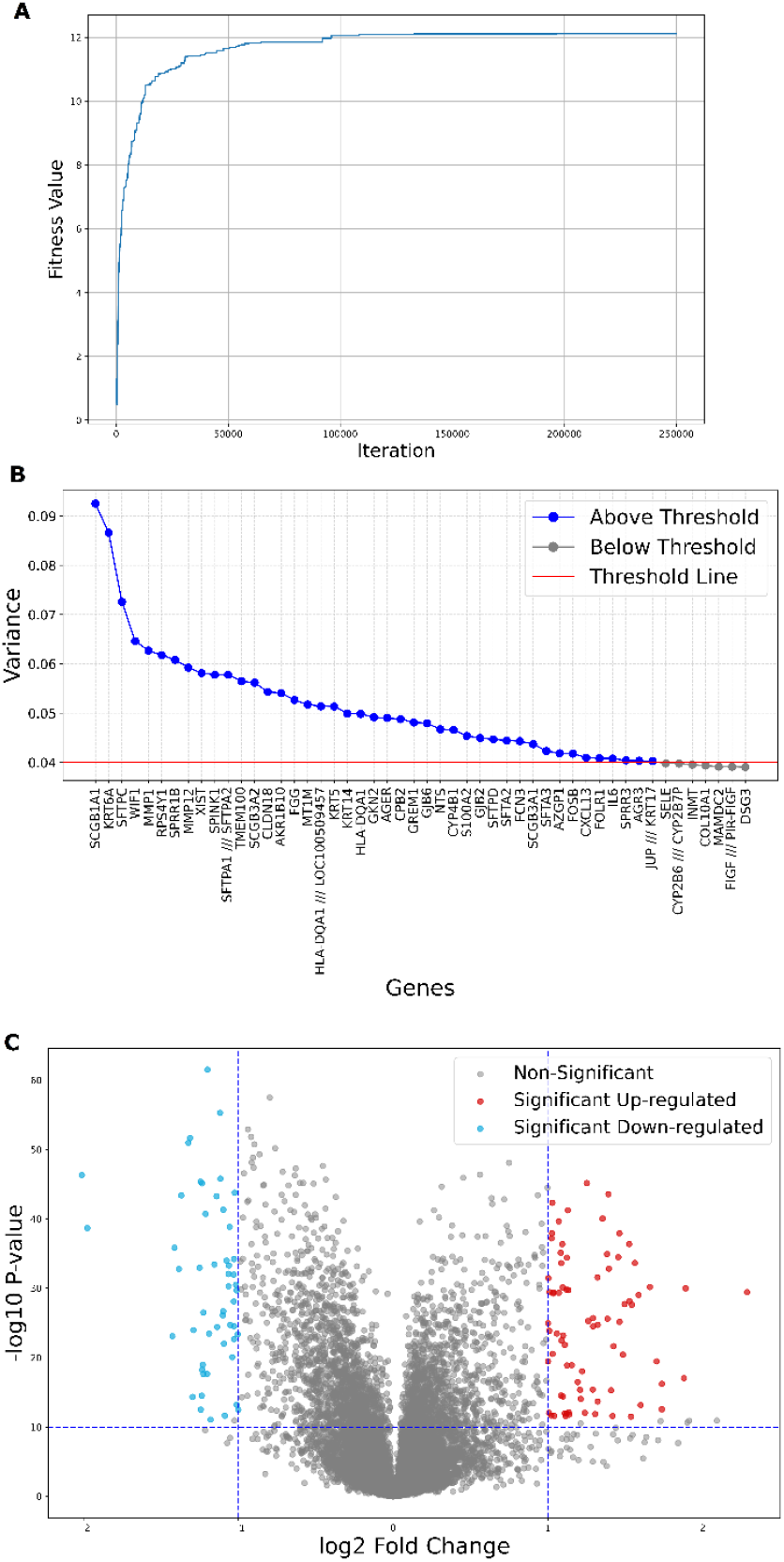
Gene selection strategy for identifying LC–associated genes. The figure illustrates the application of three complementary gene selection approaches—variance-based screening, simulated annealing–based selection, and volcano plot analysis—and their intersection to obtain a robust set of significant genes.

In order to achieve a balance between exploration and convergence, the process was carried out for 2,50,000 iterations with the annealing temperature progressively lowered using a cooling rate of 0.95. The total of the absolute mean expression differences between healthy and diseased samples throughout the chosen gene subset is the objective (fitness) function of DGESA. Instead of depending on the effects of individual genes, this approach guarantees that the chosen genes together maximize the discriminative power between phenotypic states. Fitness data were tracked during the optimization phase to keep an eye on convergence behavior. With 40 genes, the final DGESA solution achieves the smooth convergence of the fitness curve, confirming the stability and effectiveness of the optimization process (see Panel A of Fig. 2).

Genes with high variability (marked with blue dots) between samples are chosen using this method, which frequently indicates underlying biological heterogeneity. Through trial-based tuning, an empirical variance threshold of 0.04 was adopted (see Panel B of Fig. 2) to guarantee non-zero overlap between the techniques and to preserve comparability with the DGESA-selected gene set. 43 genes were chosen using this threshold, which nearly matched the size of the DGESA output. These genes exhibit highly variable expression profiles that may have biological significance.

Panel C of Fig. 2 depicts the results of the volcano plot–based gene selection method, which integrates both statistical significance and effect size. Genes were selected using stringent dual criteria: an absolute log fold change (|logFC|) threshold of 1 and a highly conservative significance threshold of −log10(P.Value) ≥ 10 (equivalent to p ≤ 1×10^−10^). Under these conditions, 130 genes were identified. While this method yields a broader gene set due to its gene-wise hypothesis-testing nature, it effectively captures genes with strong differential expression patterns.

The gene selection process applied to the integrated datasets involved three methodologies—variance-based screening, DGESA, and volcano plot analysis—which identified 40, 43, and 130 genes, respectively. See Supplementary Tables 3, 4, and 5 for the gene symbols identified by the three respective methods. The overlap among these gene sets is illustrated in the Venn diagram in Panel A of Fig. 3 and summarized in Table 1. The Venn diagram indicates that 19 genes are shared by the variance-based selection and the simulated annealing-based approach. Twenty genes in all are shared by the volcano plot and simulated annealing methods. The variance-based and volcano plot approaches share thirteen genes. Crucially, all three gene selection algorithms consistently identify 11 genes, suggesting a strong and method-independent gene subset.

**Fig. 3.**
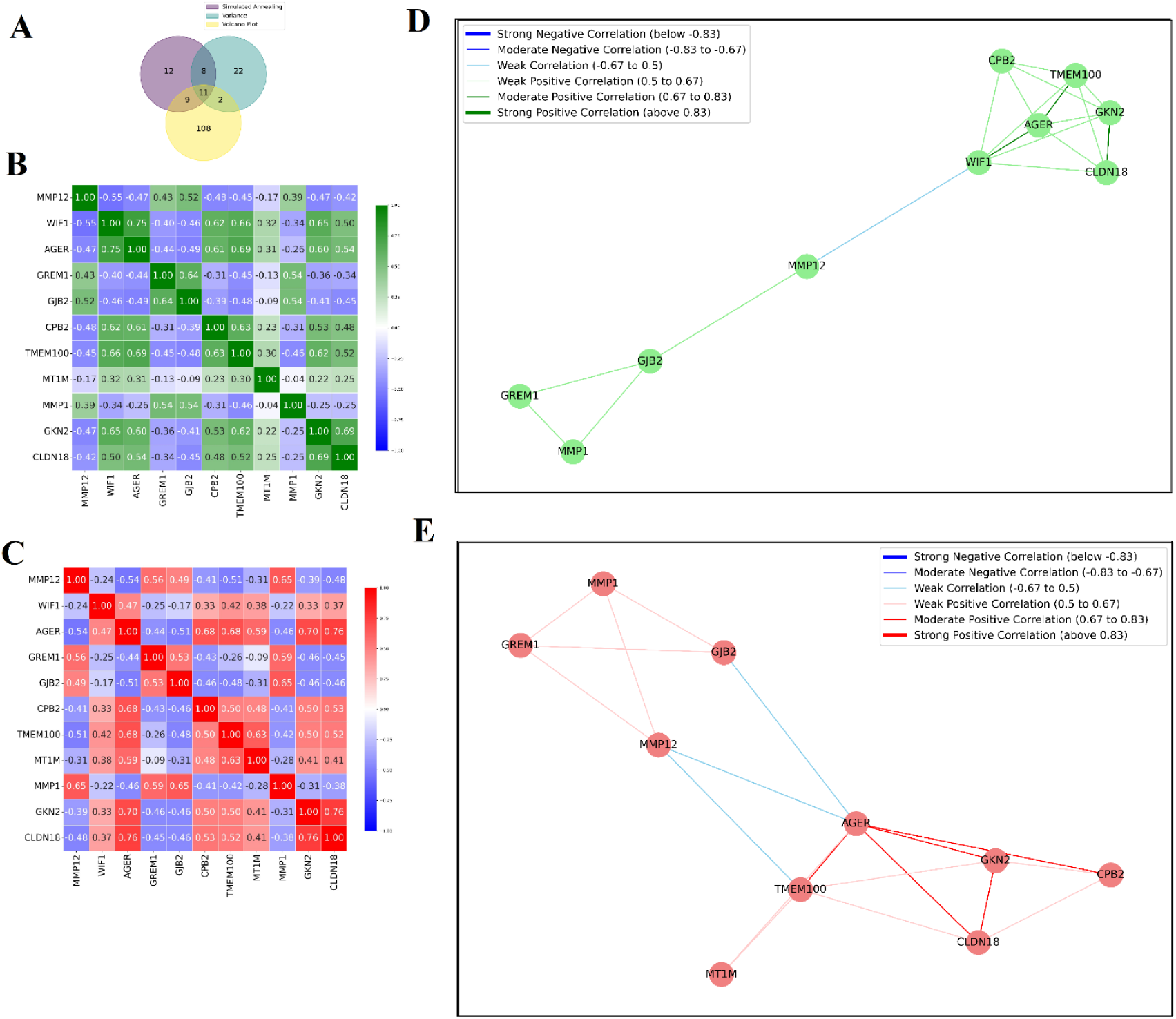
Gene selection overlap, expression patterns, and co-expression analysis. Panel A shows the overlap among genes selected by different gene selection methods using a Venn diagram. Panels B and C depict the expression heatmaps of the final 11 selected genes in healthy and diseased states, respectively. Panels D and E illustrate the corresponding co-expression networks, highlighting shifts in gene–gene association patterns between the two physiological states.

The correlation heatmaps provide a comprehensive visual representation of the 11 identified genes from the integrated dataset, illustrating their pairwise expression correlations under normal and LC conditions (Panels B and C of Fig. 3, respectively).

Co-expression-Derived GRNs were constructed for both healthy and LC states using the 11 identified genes (see Panels D and E of Fig. 3, respectively). The networks are designed based on co-expression relationships derived from correlation analysis and a predefined threshold of 0.3. Each pair of genes in the networks is connected by edges, where the edge properties (color and thickness) represent the strength and direction of their correlation. For negative correlations, edges are colored blue. Thick blue edges indicate strong negative correlations (numbers between −1 and −0.75), whereas thin blue edges indicate moderate negative correlations (values between −0.75 and −0.5). The color pale sea green with thin margins indicates weaker negative associations (values between −0.5 and −0.3). Positive correlations are visualized distinctly for healthy and disease states. In the LC network, weak positive correlations (values from 0.3 to 0.5) are represented by light coral with thin edges, while moderate positive correlations (values from 0.5 to 0.75) use red with thin edges.

Strong positive correlations (values from 0.75 to 1) are shown as Indian red with thick edges. Light green with thin margins indicates weak positive correlations (numbers between 0.3 and 0.5) in the healthy network, whereas green with thin edges indicates moderate positive correlations (values between 0.5 and 0.75). Thick green edges indicate strong positive correlations (values between 0.75 and 1). This systematic construction (see Panels D and E of Fig. 3) allows for a clear comparison of co-expression patterns between the normal and LC states, highlighting how gene interactions shift with disease progression.

### D. FRF–Based GRN Inference and Differential Network Analysis

The FRF-based GRN analysis was performed on the final set of 11 genes (MMP12 [35], WIF1 [36], AGER [37], GREM1 [38], GJB2 [39], CPB2 [40], TMEM100 [41], MT1M [42], MMP1 [43], GKN2 [44], and CLDN18) [45] derived from the overlap of the three gene selection strategies. The integrated expression dataset was first divided into healthy (Control) and diseased (Tumor) samples to enable condition-specific regulatory inference. All analyses were conducted with a fixed random seed (42) to ensure reproducibility.

For each target gene, a Random Forest regressor was trained using the remaining genes as candidate regulators. Each model employed 1000 decision trees, providing stable and robust estimates of regulatory importance while minimizing variance in feature importance scores. This process was repeated independently for all target genes in both healthy and diseased conditions, resulting in a comprehensive collection of directed regulators–target relationships.

A dynamic edge-weight threshold was estimated directly from the data. Feature importance values obtained from all Random Forest models across both conditions were pooled, and zero-valued importances were discarded to remove non-informative interactions. The final threshold was set at the 80th percentile of the combined importance distribution. This percentile-based selection ensured that only the strongest regulatory influences were retained, while maintaining a consistent sparsification criterion for both healthy and diseased networks.

Using this threshold, two directed networks were constructed per condition: a full FRF-GRN containing all inferred edges and a filtered FRF-GRN retaining only edges with weights exceeding the threshold. The filtered networks (healthy and LC) exhibited substantially reduced connectivity, emphasizing dominant regulatory interactions and improving interpretability (see panels A and B, Fig. 4). The regulatory weight matrices derived from GRN analysis are visually represented clearly and understandably using heatmaps. Regulatory weights are standardized in the range [0, 1], where values near 0 imply weak or insignificant interactions and values close to 1 suggest a high regulatory effect. Each heatmap utilizes a color gradient to represent the degree of regulatory influence between regulator–target gene pairs. Rapid identification of dominant regulators, strongly impacted targets, and global connectivity patterns within the network is made possible by the arrangement of regulator genes down one axis and target genes along the other. Regulatory rewiring and condition-specific changes in gene–gene interactions linked to biomarker transitions are further highlighted by comparing heatmaps for healthy and LC states (see panels C and D, Fig. 4).

**Fig. 4.**
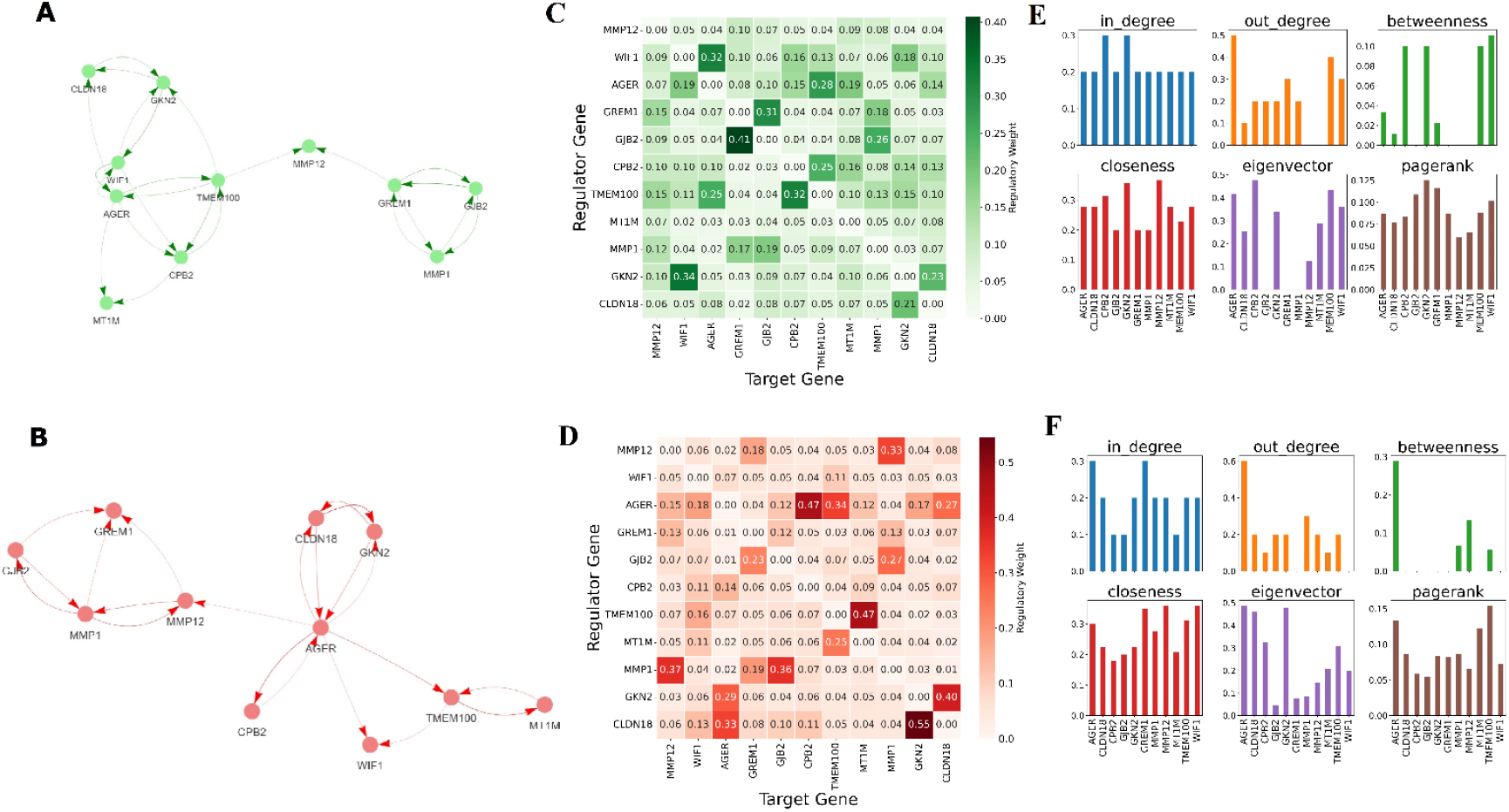
FRF–modeled GRNs in healthy and diseased states. Panels A and B show the heatmaps of Random Forest–derived regulatory weights (range [0, 1]) for healthy and diseased conditions, respectively. Panels C and D depict the corresponding thresholded FRF-GRNs, highlighting strong directed regulatory interactions retained above the data-driven importance threshold. Panels E and F present the centrality measures computed from the inferred networks, illustrating condition-specific shifts in regulatory influence and network topology between healthy and diseased states.

Centrality measures were computed on the directed Random Forest–based GRNs inferred for healthy and diseased states. Using the weighted adjacency matrices, in-degree and out-degree centralities quantified incoming and outgoing regulatory influence, respectively. Betweenness, closeness, eigenvector centrality, and PageRank were then calculated to capture control, information flow, and global importance of genes within each network. Centrality values were obtained separately for full and threshold-filtered GRNs and compared across conditions. This analysis revealed distinct shifts in regulatory influence between healthy and LC states, enabling the identification of genes with altered network roles during disease progression (see panels C and D, Fig. 4).

CLDN18, CPB2, GJB2, and MT1M are among the 11 identified genes that show pronounced alterations in their immediate regulator–target relationships in the LC state compared to the healthy state. For instance, in the healthy condition, CLDN18 is regulated by AGER and GKN2 and targets GKN2. In the LC state, while its existing regulatory links are largely preserved, AGER additionally emerges as a new target, indicating disease-specific rewiring (see Panel A of Fig. 5). CPB2 is regulated by AGER, WIF1, and TMEM100 and targets MT1M and TMEM100 in the healthy state; however, in LC, AGER remains the sole interacting gene, acting as both regulator and target (see Panel B of Fig. 5). For GJB2, GREM1 and MMP1 function as both regulators and targets under healthy conditions; in LC, GREM1 loses its regulatory role but continues as a target, whereas MMP1 retains both roles (see Panel C of Fig. 5). Similarly, MT1M is regulated by AGER and CPB2 in the healthy state, but in LC both interactions are lost, and TMEM100 emerges as a new regulator–target partner (see Panel D of Fig. 5). These examples (Fig. 5) illustrate localized regulatory rewiring, with additional gene-specific shifts detailed in the Supplement Table 6.

**Fig. 5.**
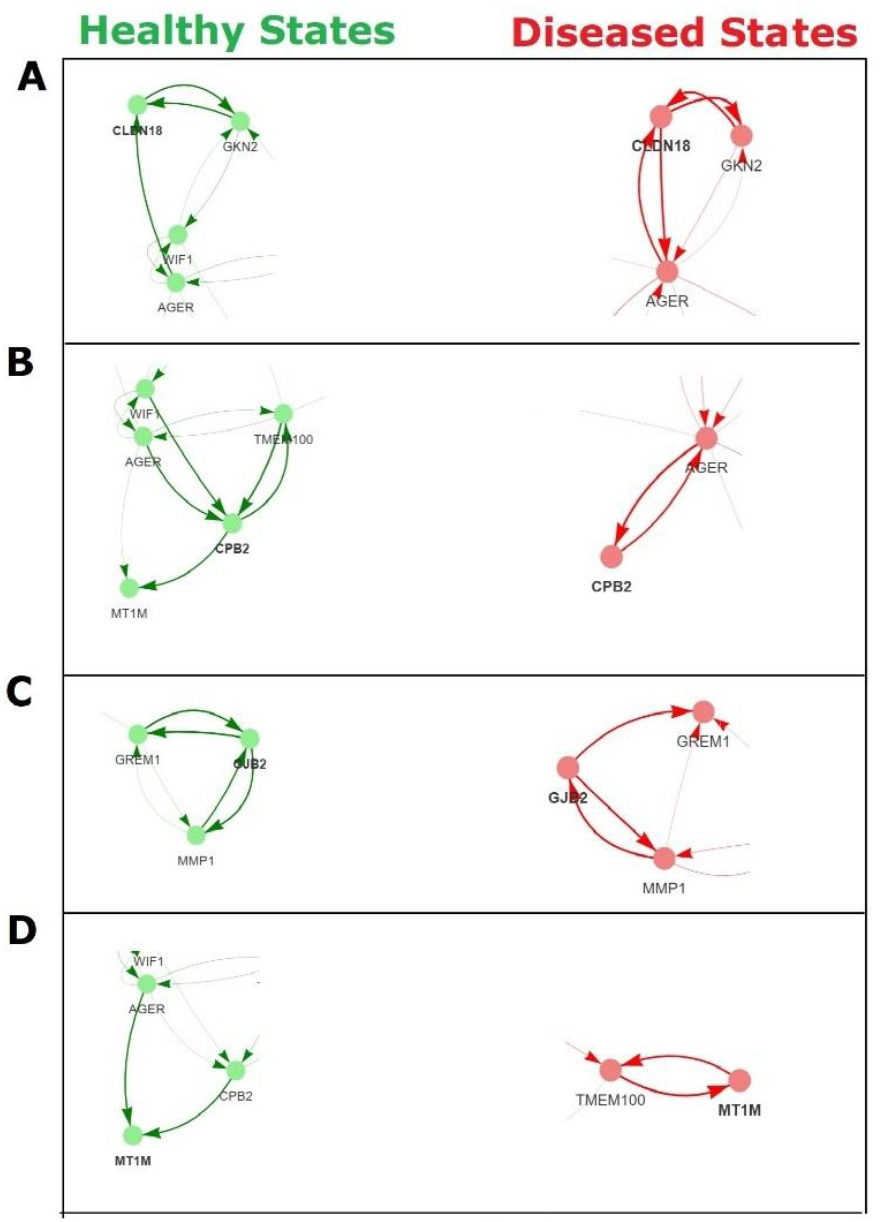
FRF–inferred GRNs illustrating regulator–target rewiring between healthy and LC conditions. The figure highlights disease-specific gains, losses, and shifts in direct regulatory interactions among the 11 identified genes, including CLDN18, CPB2, GJB2, and MT1M.

### E. GDRF–Based GRN Inference and Differential Network Analysis

Using the proposed GDRF framework, we inferred the diseased-state GRN by integrating regulator-oriented and target-oriented Random Forest outputs.

Two independently inferred interaction files—Regulator→Target and Target→Regulator—were merged to construct a unified directed network. Gene identifiers were standardized (uppercase, whitespace-trimmed), and regulatory weights were converted to numeric values, with missing entries set to zero and all weights rounded to four decimal places. Only positive-weight interactions (weight > 0) were retained, ensuring that the final network captured confident regulatory relationships.

The adjacency matrix was created by explicitly focusing on the 11 identified genes. In the target-oriented phase, each of these genes was considered a target, and a Random Forest model was used to find up to three regulators from the whole genome. In the regulator-oriented phase, the same 11 genes served as potential regulators, and for each, up to three targets were identified from all genes. All unique genes involved—including the 11 identified genes, their inferred regulators, and their inferred targets—were combined to form the node set. A weighted, directed adjacency matrix was then built based on the inferred regulatory relationships, with rows representing targets and columns representing regulators. This adjacency matrix formed the basis of the GRN. The entire process was independently performed on healthy and diseased gene expression data from the combined dataset.

The final GDRF networks for both healthy and LC conditions were modeled as directed graphs (DiGraphs) and visualized (see Panel A and B of Fig. 6 respectively) using a spring layout algorithm with a fixed seed value of 42 to ensure reproducibility. Nodes were rendered with a uniform size of 300, while edges were colored according to regulatory weights, using green for the healthy state and red for the LC condition to emphasize differences in interaction strength. All network visualizations were generated at a resolution of 300 dpi. This implementation produced stable, weighted, and directed GRNs, enabling downstream analyses such as thresholding, comparative topological assessment, and centrality-based identification of key regulatory genes.

**Fig. 6.**
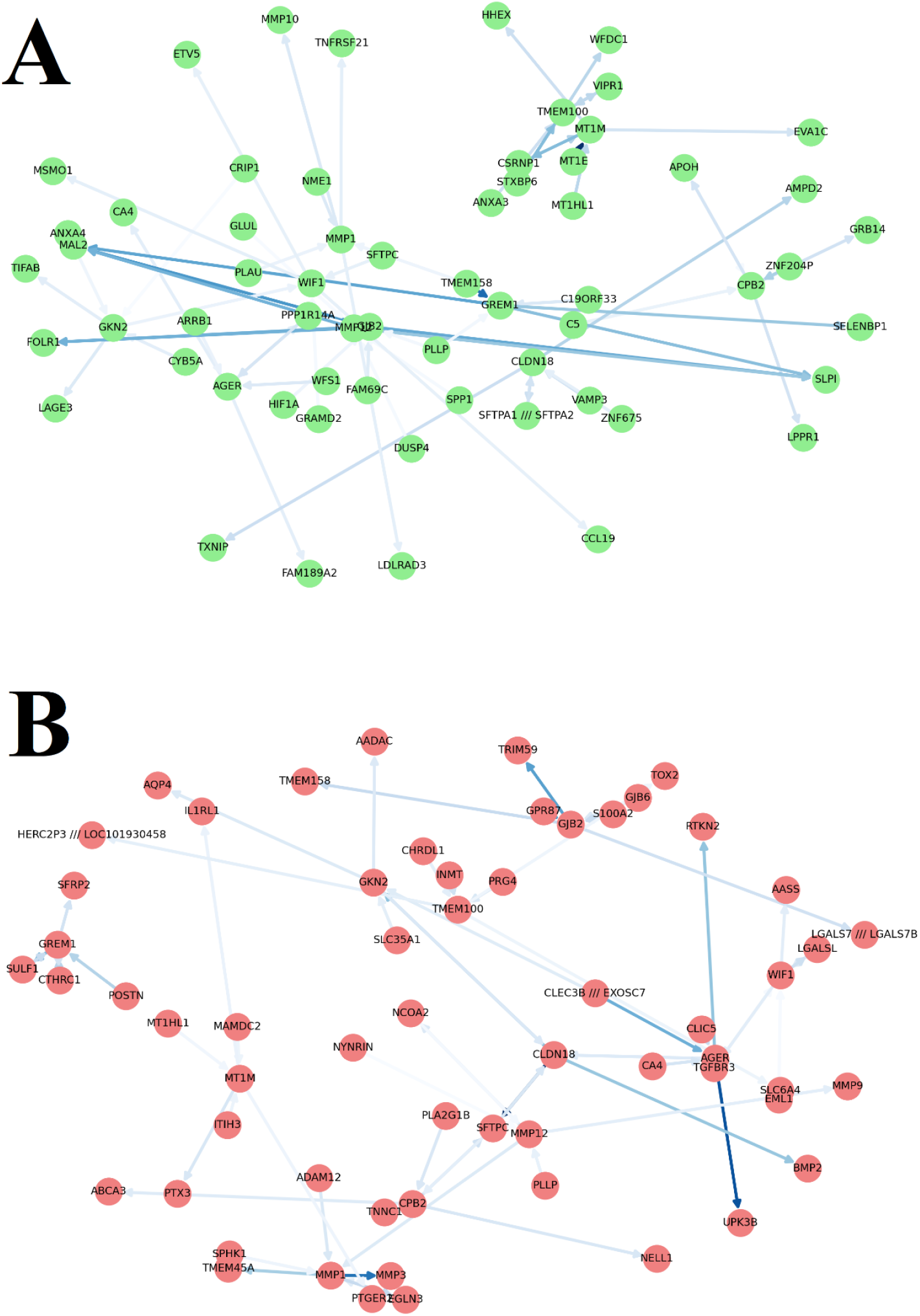
GRNs for LC and healthy samples are inferred using the GDRF architecture. The image highlights regulatory rewiring and changes in gene significance between normal and LC states, as well as condition-specific variations in directed regulator–target interactions and network structure.

To highlight important regulators in the GDRF-inferred networks, the top 10 genes for each of the three centrality measures— hub score, betweenness, and eigenvector centrality—are presented (see Panel A and B of Fig. 7 for healthy and LC conditions). Clear condition-specific regulatory rearrangement between healthy and LC states is seen based on these metrics. While GKN2 and WIF1 exhibit higher betweenness, indicating intermediary control roles, GJB2, MMP12, and GREM1 dominate hub centrality in the healthy network, indicating wide regulatory interconnectedness. TMEM100 and VIPR1 are highlighted by eigenvector centrality as significant nodes connected to other significant genes. The LC network, on the other hand, shows a clear shift, with CLDN18 becoming the dominating hub and eigenvector-central gene, followed by CPB2. Disease-specific regulatory rewiring is shown in the increased betweenness of CLDN18 and SFTPC in LC.

**Fig. 7.**
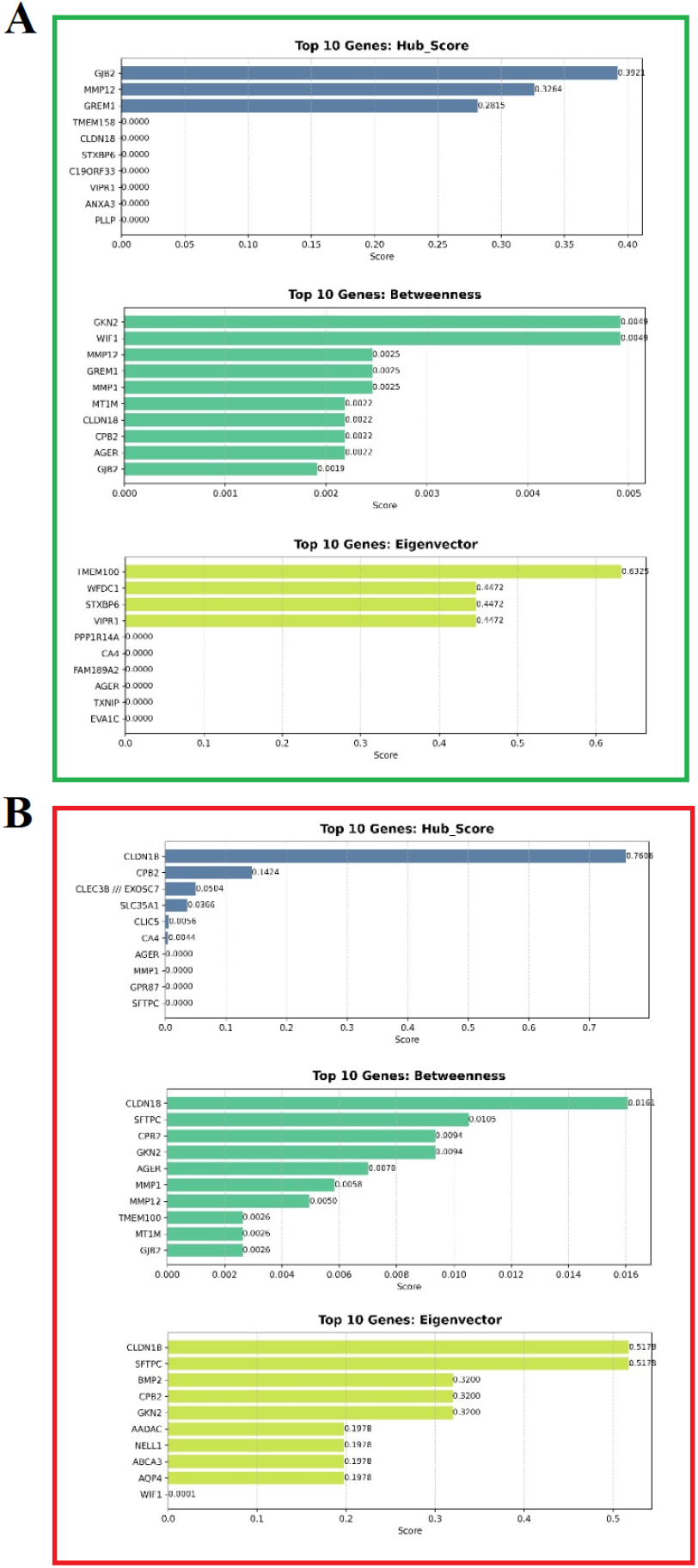
Centrality analysis of GDRF-inferred GRNs in healthy (Panel A) and LC conditions (Panel B). The figure summarizes the top-ranked genes based on hub score, betweenness, and eigenvector centrality.

## 4. Conclusion

By advancing from isolated differential expression to systemic regulation rewiring, our work aims to enhance GRN-based biomarker transition analysis, which is crucial for understanding complex diseases such as LC. A systems-level approach is crucial because disease development is driven by coordinated regulatory alterations rather than single-gene effects. This work aims to eliminate dataset-specific bias and establish a unified framework for characterizing regulatory transitions from normal to malignant states by combining two distinct LC gene expression datasets.

In order to get a trustworthy list of disease-relevant genes, a robust gene selection stage was initially implemented using variance-based screening, DGESA, and volcano plot analysis after dataset integration and normalization. To further capture regulatory changes between healthy and LC states, GRN inference was carried out at three complementary levels. To investigate global correlation shifts, co-expression networks were built. The chosen genes were then utilized to infer directed networks using a FRF technique, allowing for asymmetric regulator–target modeling with data-driven edge filtering. Lastly, by combining regulator-centric and target-centric inference, GDRF framework placed these genes inside a genome-wide regulatory context. Condition-specific regulation rearrangement was measured using centrality metrics and comparative network analysis.

Eleven strong LC-associated genes, including MMP12, WIF1, AGER, and GREM1, were found when the three screening techniques were combined. FRF-based GRNs showed substantial rewiring between healthy and LC states as well as directed, condition-specific regulatory interactions. Significant changes in the regulatory effect of genes including CLDN18, CPB2, GJB2, MT1M, and TMEM100 were shown by centrality analysis. A clear shift in network dominance from genes like GJB2 and GREM1 in healthy settings to CLDN18 and CPB2 in LC was shown by GDRF analysis, which further placed these genes within a genome-wide regulatory context. This suggests a disease-specific reorganization of control and information flow.

This work is innovative in that it uses GDRF, a dual-direction Random Forest framework to integrate local and global GRN viewpoints. In contrast to previous decision tree-based methods, this work uses Random Forests to achieve both target-centric and regulator-centric inference, combining them into a single directed network. This approach provides mechanistic insights into biomarker phase shifts in LC by enabling interpretable, weighted, and condition-aware modeling of regulatory transitions.

Although the current study concentrates on static transcriptome data, in order to improve causal interpretability, future additions might include temporal expression profiles, multi-omics data integration, and experimental validation. The suggested framework provides a generalizable method for GRN-based biomarker transition analysis since it is flexible and scalable, making it relevant to other complicated diseases and multi-condition datasets.

## Data Availability

GSE18842: https://www.ncbi.nlm.nih.gov/geo/query/acc.cgi?acc=GSE18842
GSE19188:
https://www.ncbi.nlm.nih.gov/geo/query/acc.cgi

https://www.ncbi.nlm.nih.gov/geo/query/acc.cgi

## Notes

### Competing Interest Statement

The authors have declared no competing interest.

### Funding Statement

No Funding

### Author Declarations

The study used (or will use) ONLY openly available human data at GEO, NCBI with following accession numbers: 1. GSE18842: https://www.ncbi.nlm.nih.gov/geo/query/acc.cgi?acc=GSE18842 2. GSE19188: https://www.ncbi.nlm.nih.gov/geo/query/acc.cgi

### Summary of Updates

In this updated version, we integrated two independent gene expression databases to minimize dataset-specific bias and enhance network robustness. We further introduced two Random Forest based GRN inference models: FRF, which constructs regulatory networks using only the identified candidate genes, and GRF, which infers genome-wide regulatory interactions. This dual modeling strategy enables systematic comparison between focused and global regulatory architectures, improving the reliability and biological relevance of network-based biomarker discovery.

